# ATTENTIONAL MODULATION OF NEURAL DYNAMICS IN TACTILE PERCEPTION OF COMPLEX REGIONAL PAIN SYNDROME PATIENTS

**DOI:** 10.1101/2020.10.14.20212464

**Authors:** Serena Defina, Maria Niedernhuber, Nicholas Shenker, Christopher Brown, Tristan A. Bekinschtein

## Abstract

Body perceptual disturbances are an increasingly acknowledged set of symptoms and possible clinical markers of Complex Regional Pain Syndrome (CRPS), but the neurophysiological and neurocognitive changes that underlie them are still far from being clear. We adopted a multivariate and neurodynamical approach to the analysis of EEG modulations evoked by touch to highlight differences between patients and healthy controls, between affected and unaffected side of the body, and between “passive” (i.e. no task demands and equiprobable digit stimulation) and “active” tactile processing (i.e. where a digit discrimination task was administered and spatial probability manipulated). When correct identifications are considered, an early reduction in cortical decodability (28–56 ms) distinguishes CRPS patients from healthy volunteers. However, when error trials are included in the classifier’s training, there is an unexpected increased decodability in the CRPS group compared to healthy volunteers (280–320 ms). These group differences in neural processing seemed to be driven by the affected rather than the unaffected side. We corroborated these findings with several exploratory analyses of neural representation dynamics and behavioural modelling, highlighting the need for single participant analyses. Although several limitations impacted the robustness and generalizability of these comparisons, the proposed analytical approach yielded promising insights (as well as possible biomarkers based on neural dynamics) into the relatively unexplored alterations of tactile decision-making and attentional control mechanisms in chronic CRPS.

## Introduction

Body perception is a fundamental ingredient of human conscious experience (Ehrsson *et al*., 2004; Lenggenhager *et al*., 2007; Blanke *et al*., 2015) and a strikingly relevant aspect of such experience, especially when we are in pain. A growing number of neuroimaging studies and neurocognitive models are starting to uncover the complex, multilevel, system of interactions between cortical and subcortical brain areas and afferent somatosensory information, that underpin body perception during pain (e.g., (Woo *et al*., 2017; Mouraux & Iannetti, 2018)). As it is often the case, peculiar distortions of body perception that emerge in certain chronic pain conditions, have sparked precious insights into the complexities of this important brain function. Complex Regional Pain Syndrome (CRPS) is a debilitating and poorly understood condition, in which disproportionate pain and a variable combination of other symptoms follows an apparently minor injury to one of the limbs (Bruehl, 2015; Birklein *et al*., 2018). Although this eerie “unilateral” pain condition has raised fascination and perplexity for decades, its etiology and treatment remain elusive, also because of the disarming heterogeneity of its clinical presentation (Marinus *et al*., 2011). In recent years, the observation of a disparate and peculiar set of disturbances in body perception (Lewis & Schweinhardt, 2012) (e.g., finger misperception) (Förderreuther *et al*., 2004; Kuttikat *et al*., 2017) in some of these patients has tilted the investigation of the pathophysiology of the disease towards possible (structural and functional) cortical changes in somatosensory brain regions (Swart *et al*., 2009). The initial idea, developed in analogy to other pain conditions such as phantom limb or chronic back pain (Flor *et al*., 1995; Flor *et al*., 1997; Bray & Moseley, 2011), was that misperception symptoms could result from CRPS-induced maladaptive plasticity (i.e. cortical reorganization) in the contralateral primary sensory cortex (S1) (Maihöfner *et al*., 2004; Pleger *et al*., 2004). However, the few neuroimaging studies conducted so far have led to contradicting results, spanning from a similar representation of affected and unaffected sides of the body, to an enlarged (rather than shrunk) representation of the affected limb onto S1 (Di Pietro *et al*., 2013; Di Pietro *et al*., 2015; van Velzen *et al*., 2016; Mancini *et al*., 2019; Pfannmöller *et al*., 2019).

However, as recently pointed out by some authors (Kuttikat *et al*., 2018; Brown *et al*., 2020), most work on CRPS-related body misperception (mirroring the larger branch of body perception research) has traditionally focused on early, more “physiological” components of somatosensory processing (i.e. < 50 ms; e.g., (Pleger *et al*., 2004; Lenz *et al*., 2011)), relying on experimental architectures that are more concerned with stimulation precision rather than its ecological validity. Yet, later-latency, more “cognitive” factors such as attentional modulation and perceptual decision making have long been known to modify somatosensation (Mima *et al*., 1998; Franz *et al*., 2015), including that of pain (Bantick *et al*., 2002; Chan *et al*., 2012; Clauwaert *et al*., 2020) and, the growing evidence of executive dysfunction in CRPS (Apkarian *et al*., 2004; Geha *et al*., 2008; Lee *et al*., 2015), led some to suggest that the contradictory patterns of neural representation alterations emerging from CRPS literature could be partially explained by differential cognitive and attentional engagement elicited different paradigms.

Besides these conceptual limitations, studies on CRPS-related functional plasticity are guilty of a number of further methodological issues, namely, for instance, the lack of adequate spatial resolution and accuracy of the currently available source reconstruction methods (Maihöfner *et al*., 2003; Kuttikat *et al*., 2016) and the limited sensitivity offered by average-based signal analysis techniques such as ERPs. Indeed, classical (univariate) ERP analyses by definition mask the variability across subjects in order to let similarities emerge (Luck, 2005), but individual differences in patterns of activation do exist and, especially for more heterogeneous groups of people, like CRPS patients definitely are, group contrasts are easily overwhelmed by the amount of inter-individual noise.

Multivariate pattern analysis (MVPA; or “brain decoding”) is gaining exponential popularity in the world of cognitive (and clinical) neuroscience, thanks to the promise of increased sensitivity and reduced reliance on “spatially variant” confounds such us, e.g., anatomical differences in the folding pattern of the cortex (King & Dehaene, 2014; Grootswagers *et al*., 2017). MVPA offers additional advantages when investigating the temporo-spatial dynamics of pain processing (Rosa & Seymour, 2014) and it has successfully been used to decode pain experience and sensitivity in healthy volunteers (Schulz *et al*., 2011; Tu *et al*., 2014; Lancaster *et al*., 2017). However, applications of decoding techniques to EEG signals (that are conveniently portable, cheap, and offer excellent temporal precision) have hardly been used in combination with attentional demand manipulations to study (painful) somatosensation in healthy humans, nor in any clinical setting. Hence, we aimed at investigating the influence of attentional modulation on CRPS-related perceptual alterations, by using multivariate pattern classification of early, mid-latency and late EEG activity elicited by tactile digit stimulation and recognition. We exploited MVPA methodological advantages to better reflect and leverage individual patterns of “neural information” and highlight differences between CRPS patients and healthy volunteers (HV) and between the affected and unaffected side in individual patients. We did so not only during “rest” (i.e. passive stimulation) but also under increased “cognitive” demands (i.e. active condition, where spatial probability was manipulated, and a digit discrimination task was administered). As per our hypotheses (https://osf.io/rmhsb), we expected:

H.1. A main effect of group (CRPS < HV) on mean classifier performance in distinguishing the five fingers from each other (i.e. *all-vs-all* decoding).
H.2. An interaction of group by side (left and right) on *all-vs-all* finger classifier accuracy, in the direction of smaller difference between hands in the HV group.
H.3. A main effect of side (affected < unaffected) on classifier performance for brain activity elicited by stimulation of the thumb (D1) vs. the little finger (D5) (i.e. *1-vs-5* decoding), across conditions (passive and active stimulation), in any of the CRPS patients.
H.4. An interaction of condition by side-affected on *1-vs-5* decoding accuracy, such that, in the active but not in the passive condition the performance in the affected hand will be comparable to that of the healthy (unaffected) hand. In other words, the enhanced attentional demand induced by the task would compensate for poor sensory processing on the affected side, thereby reducing the decodability gap between sides in the CRPS group.

Despite their mostly descriptive and exploratory nature, we believe our results shed important insight into the possible neurophysiological and neurocognitive basis of tactile misperceptions reported in CRPS, opening a window for a conceptually and methodologically new approach to this field of investigation.

## Materials and methods

### Participants

Our sample included 13 patients (11 females, mean(range) age = 46.8(30-63) years) who were diagnosed with unilateral upper or lower limb CRPS according to modified Budapest Research Criteria (Harden *et al*., 2007) and 13 age and sex matched healthy volunteers (11 females, mean(range) age = 45.0(28-63) years). All participants (patients and healthy volunteers) were right-handed. Exclusion criteria were: previous or current diagnosis of peripheral neuropathy, stroke, transient ischemic attack, multiple sclerosis, malignancy or seizure disorder. Table 1 provides a summary of the sample. An overview of the clinical features of all included CRPS patients is reported in Supplementary Table 1.

**Table 1.**
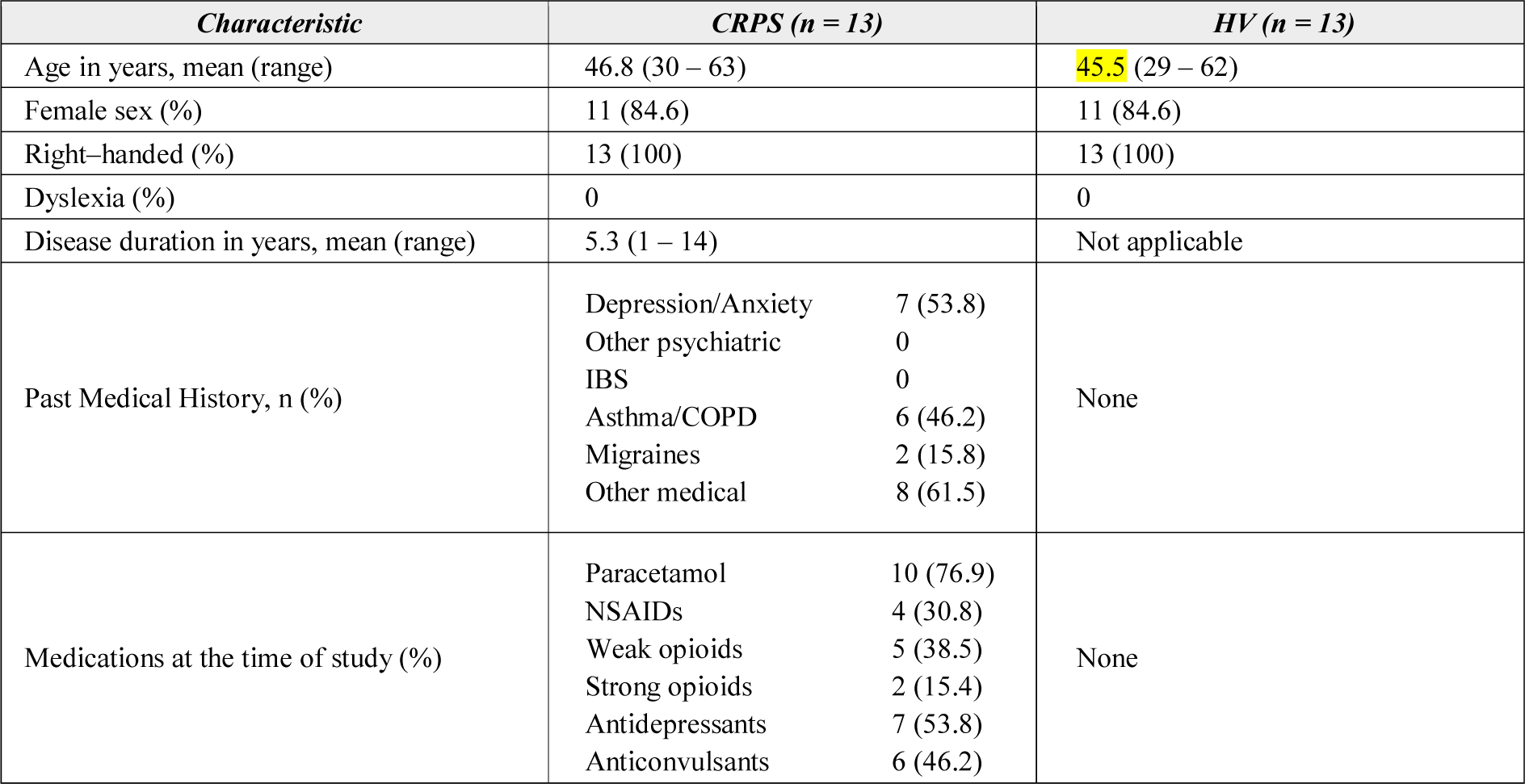
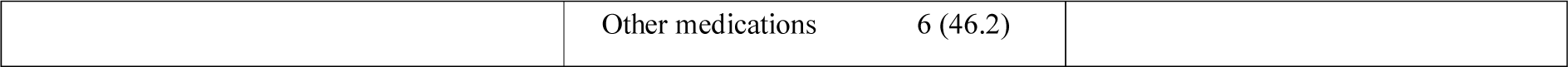
Summary of baseline characteristics of study participants including healthy volunteers (HV) and CRPS patients’ including clinical features.

### Ethical standards

All participants gave written and informed consent to take part in the study and were remunerated for their time. The study was approved by the Research Ethics Committee of East of England-South Cambridge (REC Ref No: 12/EE/0305) and it conforms with the World Medical Association Declaration of Helsinki (2013). All possible measures were taken to minimize pain or discomfort in all participants, with a special attention to the CRPS group. All patients were advised to report immediately if the sensation was uncomfortable or painful and the fingertips were also checked after each session for any redness of the skin. After every block of sensory stimulation, a break of at least 10 seconds (in the passive condition) or a few minutes (in the active condition) was offered to improve participants’ comfort.

### Study design

Two custom-made handboxes (one for each hand) were calibrated to deliver soft touch stimuli with the same force to the fingertips of each digit. The experiment, as depicted in *Figure 1*, consisted of 4 blocks of passive stimulation (i.e. passive condition), followed by 8 blocks of finger recognition task (i.e. active condition). In the passive condition, participants were instructed to sit relaxed and keep their eyes closed and head still as much as possible. In each passive block, 50 touches were delivered in random order to each finger of one hand. Left and right hand were alternated between blocks, for a total of 500 trials per hand (100 for each finger). In the active condition, participants placed one hand on the handbox, with their eyes still closed and a relaxed but still posture; again, hands were alternated between blocks. This time, participants were asked to respond to the stimulation by saying out loud the number corresponding to the stimulated finger (i.e. thumb = 1, index finger = 2, middle finger = 3, ring finger = 4 and little finger = 5) and a bottom-up manipulation of predictability was implemented, with outer fingers (D1 and D5, i.e. “expected”) being touched on 75% of all trials, whereas the probability of stimulation for central fingers (D2, D3 or D4, i.e., “rare” / “oddball” fingers) was 25%. The answer was recorded manually by the experimenter thereby triggering the next stimulus. Each active block consisted of 80 trials, for a total of 320 stimulations for each hand (120 to each thumb, 120 to each little finger, and ∼27 each for the remaining three rare fingers). All tactile stimuli had a duration of 50ms, and a fixed interval of 950 ms separated single stimulations.

**Figure 1.**
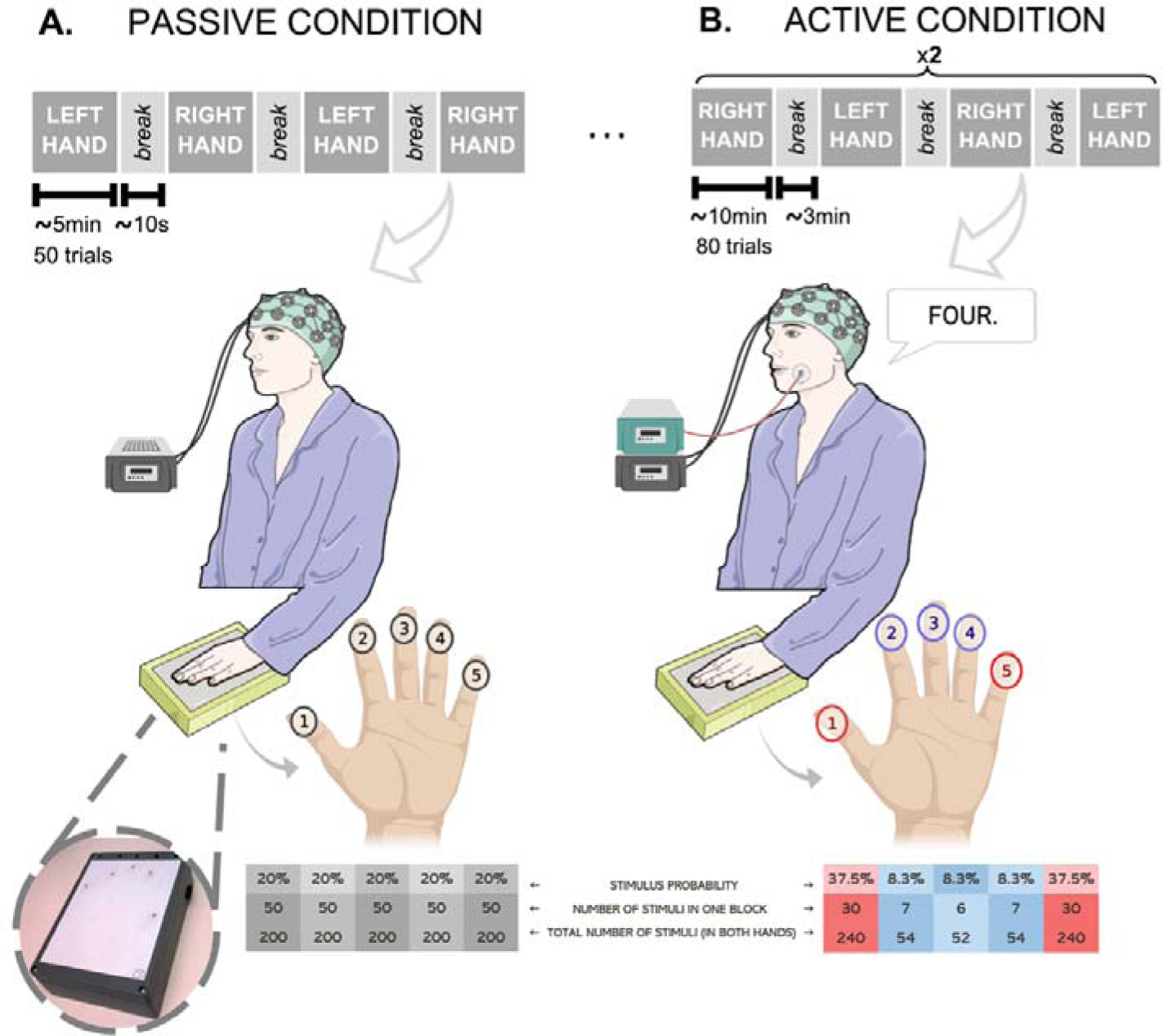
During the study visit, participants gave written informed consent and were sitting comfortably as the EEG cap was placed and prepared for recording. The handboxes (in the picture above) were then calibrated to deliver soft touch stimuli with the same force to the fingertips of each digit and soft ear plugs were provided to reduce any external auditory input and help participants focusing on the tactile stimuli. **(A)** In the passive condition the order of digit stimulation was randomised within each block (on each hand in turn). Touches were delivered at a predefined frequency of 1Hz (duration: 50ms; ISI: 950ms). Each of the 4 (∼5 minutes-long) blocks contained 250 trials (50 stimulations per digit), for a total of 1000 trials (500 per hand and 100 per individual fingers) in ∼20 minutes. A 10 seconds break was offered after every block to improve participant’s comfort. **(B)** In the active condition, digit stimulation was pseudo-randomised in each hand / block, with D1 and D5 receiving 30 stimuli each (probability = 37.5%), for a total of 240 touches each across the two hands (120 on the left and 120 on the right). The inner digits (D2, D3 and D4) shared the remaining 20 trials of each block (probability = 8.3%), reaching ∼54 stimuli each across the two hands (∼27 on each hand). Hence, of the total 640 active trials, 480 were dedicated to “expected” fingers (D1 and D5) and 160 to the “rare” central fingers (D2-D4). A microphone attached to the EMG recording was used to capture the participant’s reaction time (resulting from the subtraction of the stimulus onset time from the start of the voice deflection on the EMG). The answer was recorded manually by the experimenter by typing the number on the computer and triggering the next stimulus. A maximum time lock was set on three seconds per trial.

### EEG acquisition and preprocessing

During both conditions, 128-channel high-density EEG data were collected using the Net Amps 300 amplifier (Electrical Geodesics Inc., Oregon, USA), at the sampling frequency of 250 Hz. Raw data were pre-processed in MATLAB using the EEGLAB toolbox (Delorme & Makeig, 2004) and in-house scripts. Channels on the neck, cheeks and forehead were removed, leaving 92 retained channels for further analysis. EEG data were filtered between 0.5 and 30Hz and epoched between - 200ms and 800ms relative to stimulation onset. Artefacts originating from eye blinks, eye or muscle movements and electrical interference were identified and removed by means of visual inspection and independent component analysis (ICA). After the interpolation of rejected channels, data were re-referenced to the average and baseline-corrected relative to a 200ms interval before the stimulation. For each participant, single-subject ERP plots were examined to ensure data quality.

### MVPA and Statistical Analysis

Multivariate Pattern Analysis (MVPA) on EEG data was implemented using MATLAB-based ADAM toolbox (Fahrenfort *et al*., 2018). We employed a Linear Discriminant Analysis (LDA) algorithm to perform decoding (Grootswagers *et al*., 2017) and a 10-fold stratified cross-validation method was implemented to prevent overfitting. Hence, we ran the classification 10 times; each time the LDA model was fit (or “trained”) on 9/10 of the total number of trials, leaving the remaining 10% for testing. The procedure was then repeated for each time point in the trial.

To improve the interpretability of the active condition decoding output, we conducted three sets of analyses including: *a)* only correct identification trials, *b)* the full set of available active trials (including error trials) and *c)* only trials where response was registered later than 500 ms after stimulus presentation. Imbalances within and between classes resulting from noisy trial rejection and/or experimental manipulations were corrected by means of undersampling.

### Healthy super-subject analysis

In order to select the time-window(s) of relevant processing, brain decoding was first performed at a “super-subject” level for both passive and active (i.e., full set) conditions. This time-window identification process entailed four analytical steps. First, a super-subject was built by “stacking” together all trials from all healthy volunteers, regardless of who performed them. The resulting “super-block” was then analysed as a single subject dataset, following the procedure described above (step 2). For this analytical step, a “*one-vs-all*” classification strategy was adopted, meaning that classifier performance was calculated for each individual finger (against all others), e.g., discriminating “thumb” vs. “not-thumb”, where the “not-thumb” class consisted of all trials stimulating index, middle, ring or little finger. The five time-courses obtained in step 2 were then averaged to yield a single time-wise mean performance output (step 3). Finally, to determine the time-window(s) of interest for subsequent analyses, we applied cluster-based permutation testing on the mean time-course obtained in step 3, using a significance level of 0.05. This procedure reduces the number of statistical tests (i.e., multiple comparison problem) by using temporal clusters rather than individual samples as relevant units for p-value computations (Pernet *et al*., 2015). After computing sample-wise comparisons of measured performance to chance levels (i.e. 137 tests, for a window of 550 ms and a sampling frequency of 250Hz), we determined the number and size of reliable clusters, defined as temporally contiguous reliable samples (p < 0.05). We then permuted the classes labels, randomly assigning trials to e.g., “thumb” or “not-thumb”, re-tested discrimination performance vs. chance, and computed new cluster sizes. We repeated the procedure 1000 times and counted how often a reliable cluster equal or larger than the original one was observed under random permutation. The resulting cluster p-values are obtained by simply dividing the number of randomly larger clusters by 1000, and they reflect the probability that a temporal cluster of the observed size (i.e., number of consecutive timepoints where performance is reliably above chance) occurs by chance alone (Maris & Oostenveld, 2007; Pernet *et al*., 2015).

### Group-level analysis

For the “*all-vs-all*” analysis, a multi-class LDA was trained to assign the input brain pattern to one of five finger classes (D1, D2, D3, D4 and D5). To quantify the decoding performance, Classification Accuracy (CA) was calculated per class, by averaging across the 10 folds (at each time-point), then averaged across classes and compared to the random classification rate (RCR), defined as the reciprocal of the number of classes (i.e. 20%). Reliable temporal clusters were identified using cluster-based permutation tests (Maris & Oostenveld, 2007; Pernet *et al*., 2015).

### Individual-level analysis

For the “*1-vs-5*” analysis, a linear classifier was fit to the trials corresponding to stimulation of the thumb (D1) and little finger (D5) in both conditions and hands of the CRPS group. Classifier performance was measured using Area Under the Curve (AUC), a nonparametric criterion-free measure of generalization, that is commonly recommended over CA for problems with two possible classes (Grootswagers *et al*., 2017). In summary, each prediction (i.e. time-point) can be considered as an implicit instance of a confusion matrix defined by a true positive rate (TPR; e.g. how many 5s are labelled as 5s) and a false positive rate (FPR; e.g. how many 5s are labelled as 1s). The trade-off between TPR and FPR can be represented in space (Receiver Operating Characteristic or ROC space) by a curve, representing all combinations of TPR and FPR that would yield equal sensitivity / specificity. The area under this curve (AUC) serves as a measure of classification performance (sensitivity / specificity trade-off).

### Statistical analysis

To assess statistical significance (hypothesis H.1 and H.2), the selected time-windows of “*all-vs-all*” decoding output for the two groups and sides were compared using a Linear Mixed Model with “side” (affected/non-dominant vs unaffected/dominant), “group” (CRPS vs HV) and their interaction estimated as fixed effects, whereas the intercept values were allowed to vary across subjects (i.e., random coefficient). Because the difference between active and passive activations became evident in later stages of processing, we examined it by adding a main effect of condition among the fixed coefficients of the model. Finally, to test hypothesis H.3 and H.4, each patients’ data was compared in a two-way repeated-measures ANOVA featuring side (affected and unaffected) and condition (passive and active) as main factors. Statistical analysis was implemented in R and Jamovi (Şahin & Aybek, 2019) and a p-value <.05 was considered statistically reliable.

### Exploratory analysis

#### Behavioral performance

To explore whether the trends detected by our analysis would mirror any differences in the actual behavioral choices of our participants in the finger recognition task, we calculated *d’* (d-prime) and bias (*c*) scores by opposing “expected” (D1 and D5) and “rare” (D2-D4) digits. Originally borrowed from Signal Detection Theory (SDT), *d’* and *c* are meant to decompose response behavior in order to capture the individual’s sensitivity (i.e., the ability to distinguish the two alternative sensory signals) and his/her “decision criterion”, providing an indication of any “preference” for one or the other answer (Wickens, 2002; Macmillan & Creelman, 2004; Bang & Rahnev, 2017). In our case, *c* is expected to reflect the effect of expectation induced by digit probability manipulation. In other words, an “optimal” decision criterion would be slightly displaced towards negative values, as responses “1” and “5” are more probably correct and therefore “preferred” over “2”, “3” or “4”. The same model used for group-level analysis of classifier output was adopted to predict both these scores (i.e., a Linear Mixed Model with side, group and their interaction estimated as fixed effects, whereas the intercept of each subject as a random coefficient).

#### Temporal Generalization Analysis

We also assessed the temporal stability/mutability of these finger mental representations in the two groups, conditions, and sides of the body, by applying Temporal Generalization Analysis (TGA), a decoding technique that, instead of training and testing a classifier on data from a single timepoint at the time as we described above, tests each classifier trained on time *t* using all other timepoints in the trial (King & Dehaene, 2014). If the same pattern of information (e.g., the representation of a finger) recurs at multiple time points, that pattern is deemed stable. If the generalizability is low despite above-chance decoding, then the neural information that supports such decoding is likely not as stable but rather it evolves dynamically across time.

#### Topographical Maps

Finally, with the intent to better understand the pattern of neural activity that may give rise to classification performance, i.e. in space rather than in time, we derived time-specific topographical scalp maps from decoder output. This procedure was implemented using the ADAM toolbox (Fahrenfort *et al*., 2018) and performed as follows: the diagonal classifier weight vectors resulting from the main (Backward Decoding Model, or BDM) analysis were first multiplied by the data covariance matrix. These vectors contain one weight value for every electrode in the set and they are time-point specific. The multiplication effectively transforms the BDM vectors into the output of a forward model (Haufe *et al*., 2014). The resulting activation patterns can be now interpreted as neural sources (i.e. neural activation that underlies the decoding result) and plotted onto a topographical map of the scalp that is equivalent to those obtained from classic univariate analyses. Reliable spatial clusters (i.e. contiguous electrodes with p < 0.05) were computed using permutation testing, in a similar manner to the procedure described above (Maris & Oostenveld, 2007).

All the scripts used for the neural and behavioral analyses and for the displaying of results can be found at https://github.com/SereDef/CRPS-decoding-behavioral-modeling.

## Results

### Behavioral performance

Behavioural accuracy and Reaction Times (RTs) for digit identification (active task), are reported for both hands in every participant, and for individual fingers in the two groups in *Supplementary Table 2*. The mean RTs refer to correct identification trials only. In summary, behavioral performance measures show some support for our H1, as patients had overall worse mean accuracy (HV = 97%, CRPS = 82.4%; *Z*=2.28, *P*=0.011) and longer mean RTs (HV = 957.4 ms, CRPS = 1363.2 ms; *Z*=2.54, *P*=0.005) compared to healthy controls. There was some indication of worse performance on the affected side compared to the unaffected side that is specific to CRPS patients (H2), but only with respect to mean RTs (affected = 1462.2 ms, unaffected = 1264.2 ms; *Z*=2.85, *P*=0.002). A more detailed analysis of Accuracy and RT data can be found in (Kuttikat *et al*., 2018), however an important result to point out is a greater variability of CRPS patients’ performance compared to the HV group.

With respect to the exploratory analysis of the behavioral variables (represented in *Figure 2*), a reliable main effect of group (*F*_1,24_=5.83, *P*=0.024, η^2^_*p*_ =0.158) and side (*F*_1,24_=12.63, *P*=0.002, η^2^_*p*_=0.065) emerged on *d’* scores. Sensitivity scores were 0.48 SDs higher on average on the unaffected / dominant hand compared to the contralateral one, and 0.70 units lower in CRPS patients compared to the HV group. Even though the interaction effect was not reliable (*F*_1,24_=0.54, *P*=0.470, η^2^_*p*_=0.003), the difference in performance between the two hands (i.e., affected < unaffected) was higher in the CRPS group (*F*_1,24_=9.20, *P*=0.006), compared to that of healthy volunteers (*F*_1,24_=3.97, *P*=0.058). With respect to the bias (*c*) score, none of the fixed effect included in the model did yield reliable results (Group effect: *F*_1,24_=3.27, *P*=0.083 η^2^_*p*_=0.078; Side: *F*_1,24_=2.42, *P*=0.133, η^2^_*p*_=0.020; Group ⍰ Side: *F*_1,24_=0.82, *P* =0.373, η^2^_*p*_=0.006). However, as evident from *Figure 2*, the main difference between the two groups’ behavioral prerfomance was really in the accentuated spread or variability of such measures across the patients’ group compared to healthy controls.

**Figure 2.**
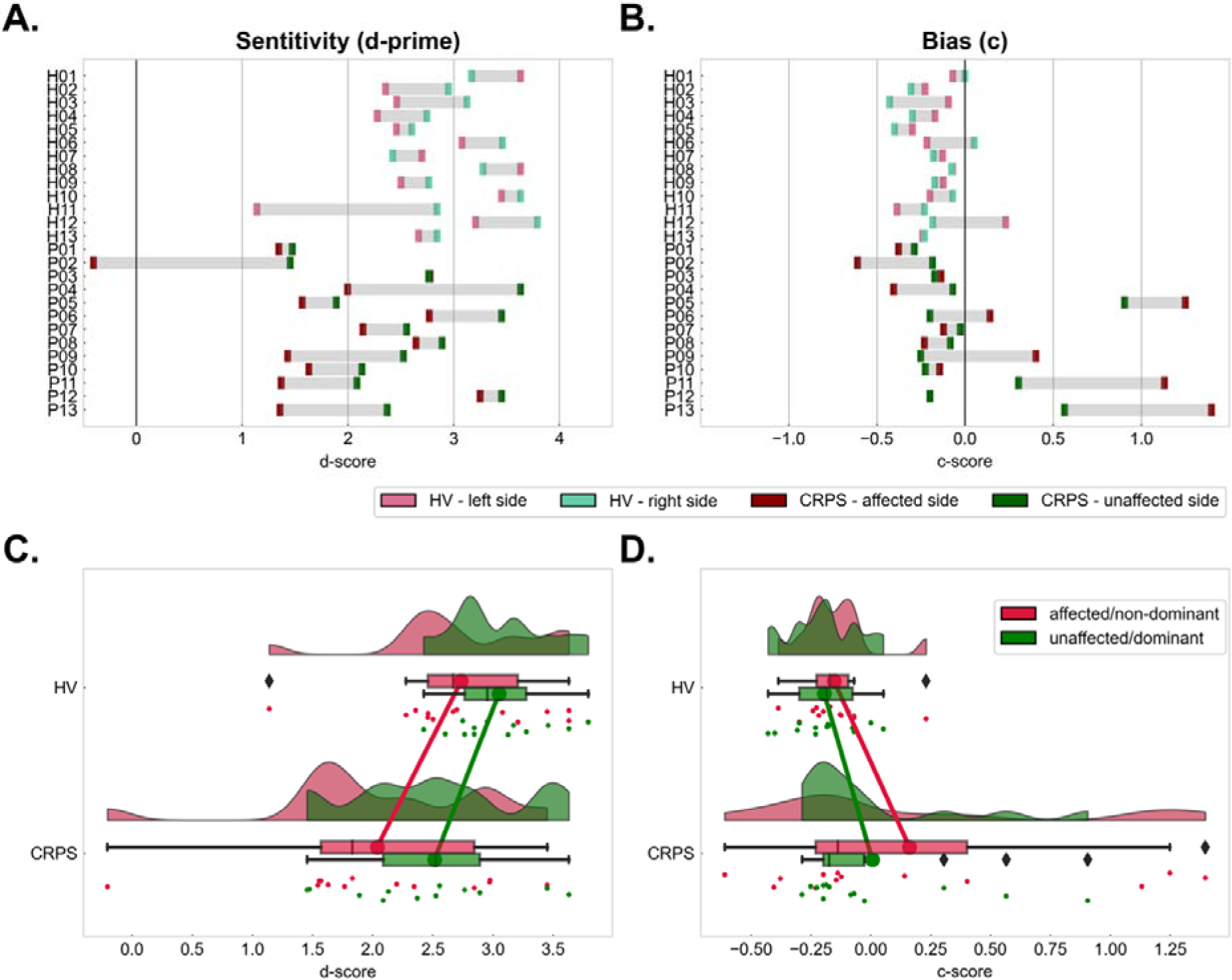
Overview of behavioral performance as measured by sensitivity (d’) and bias (c) in the two groups and sides. **(A, B)** Cleveland dot plots representing the subject-wise (A) d’ score and (B) bias scores calculated between digits 1-5 (“expected fingers”) and 2-3-4 (“rare fingers”). The affected / non-dominant side is represented in dark red for CRPS patients and in pink for HVs, while the unaffected / dominant side is represented in dark and light green (for CRPS and HV respectively). Each “row” in the plots corresponds to one participant, ordered according to participant ID and with HVs preceding CRPS patients. The individual difference between sides is highlighted by grey shadowing. **(C)** Distribution of d’ scores in the two sides of HV and CRPS groups (above and below respectively). **(D)** Distribution of c (bias) scores in the two groups and hands. The same color scheme was adopted (red for affected / non-dominant and green for unaffected / dominant) and the two transversal lines provide a representation of the side-specific mean difference between groups.

### Healthy “super-subject” results

The temporal regions of interest we defined using the mean “super-subject” classifier are illustrated in *Figure 3*. The five *one-vs-all* time-courses resulting from intermediate step 2 of the super-subject analysis are displayed in *Supplementary Figure 1*. Mean super-subject decoding performance was reliably better than chance within three temporal clusters: between 28 and 56 ms (peak: 40 ms) and between 84 and 128 ms (peak: 112 ms) after stimulus onset in the passive condition; and between 364 and 500 ms (peak: 480 ms) after stimulus onset in the active condition (Figure 3A and 3B). The same temporal patterns appeared when *all-vs-all* and a *1-vs-5* decoder were trained and tested on the same healthy “super-subject” data (Figure 3C and 3D). We therefore selected an “early” (28-56 ms), “mid” (84-128 ms) and “late” (364-500 ms) time windows for statistical testing. We cautiously decided to include a fourth “mid-late” time window, extending from 180 to 320 ms after stimulus presentation, as a large peak in decoding performance seems to be present in the active condition (Figure 3D), potentially relevant for cognitive processing. With respect to the active condition, these results refer to the full set of trials (including both error and early response trials) but were consistent across sensitivity analyses.

**Figure 3.**
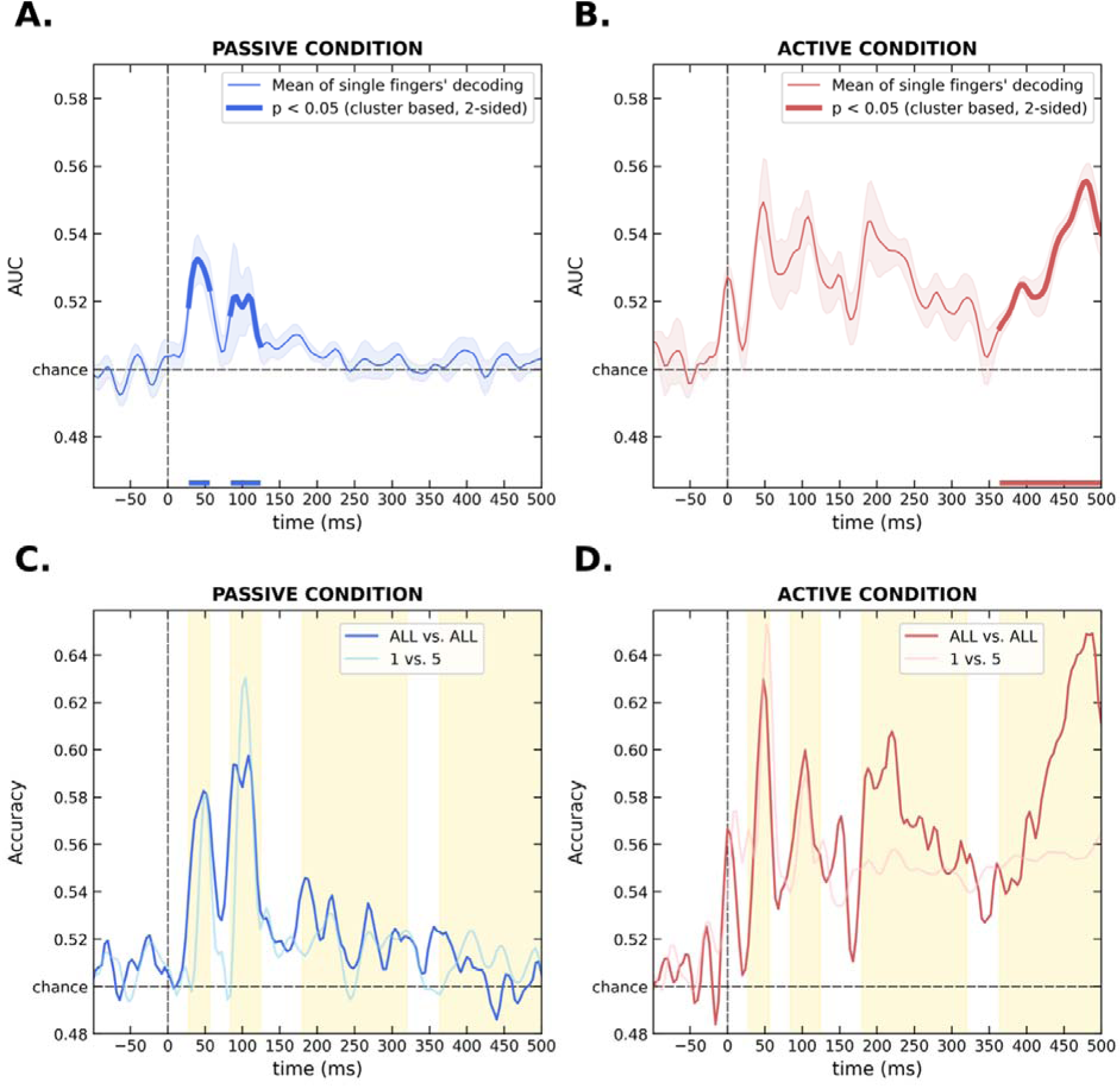
Time-courses resulting from healthy super-subject analysis. **(A, B)** The average time-course of five one-vs-all classifiers trained and tested on the healthy “super-subject” data in the passive (A) and active (B) condition. The portion of time that survived cluster-based permutation test for significance against chance is highlighted with a thicker line. **(C, D)** The decoding time-course of the *all-vs-all* and the *1-vs-5* classifiers applied to the passive (C) and active (D) super-subject data, with the four time-windows selected for further testing highlighted by the yellow shadowing.

### Group-level results

The performance (i.e., CA) time-courses of the *all-vs-all* decoder is shown in *Figure 4.* The active condition outputs (Figure 4B, D and F) are extracted from correct trials only. A comparison with the corresponding active time-courses resulting from the full set and from the >500ms RT trials only is available in *Supplementary Figure 2*. For each subject, we selected four temporal clusters of CA values (early, mid, mid-late and late time windows) and averaged CA across time within those windows. Within the same windows the peak (highest performance value) and its latency were also registered and analysed for exploration purposes. All results including F-values for each comparison, their associated p-values and effect sizes are reported in *Supplementary Table 3* (correct trials only) and *Supplementary Table 4* (all active trials).

**Figure 4.**
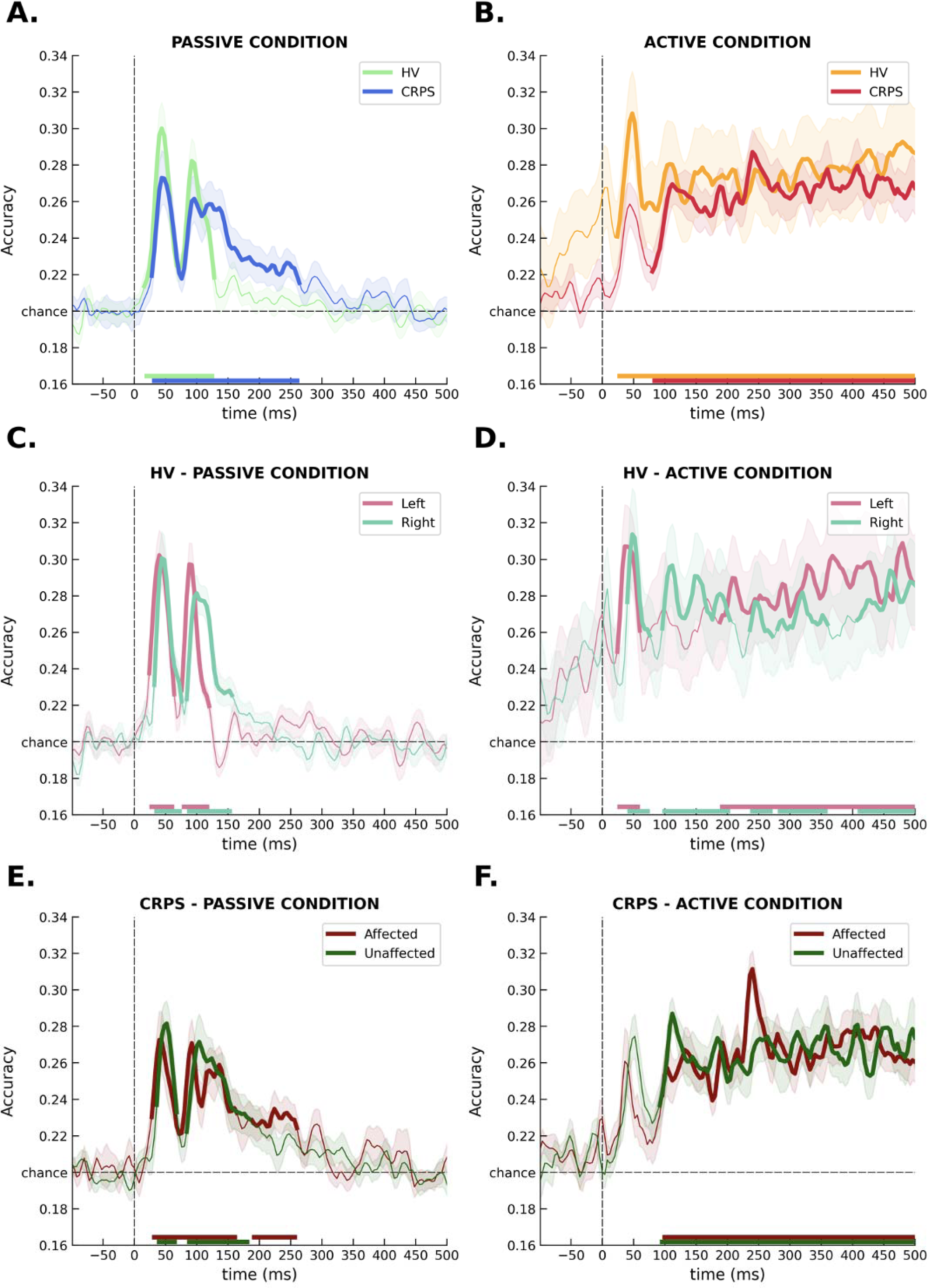
(A) Decoding performance (CA) as a function of time, resulting from training and testing two *all-vs-all* classifiers on the passive condition data from healthy volunteers (in green) and CRPS patients (in blue). **(B)** The time-course of the same two *all-vs-all* classifiers trained on the correct trials from the active condition, for healthy controls (in yellow) and CRPS patients (in red). **(C)** The passive and **(D)** active (correct trials only) time-courses of classification performance on the left (in pink) and on the right hand (in light green) in the HV group only. **(E)** The passive and **(F)** active (correct trials only) decoding performance time-course on the affected (in red) and on the unaffected side (in dark green) of the CRPS group only. In all of the above plots, the portion of time where the classifier performed reliably better than chance (RCR = 20%) was established by means of cluster-based permutation test, and highlighted with a thicker line. The output of MVPA applied to successive time points, as pictured in the six graphs, is meant to describe when and for how long the information of interest, i.e., the identity of the finger that is being touched, becomes “encoded” or “represented” explicitly in the brain (Grootswagers *et al.,* 2017).

We found evidence supporting our first hypothesis – i.e., lower mean decoding performance in the CRPS group compared to controls - in the early time window (*F*_1,25_ = 5.62, *P* = 0.026, η^2^_*p*_ = 0.067), with patients obtaining on average 3 CA units decrease compared to HV between 28 and 56 ms. Although the interaction term in the model did not reach significance, by inspecting the simple effects one can appreciate how this early group difference was driven by the affected (*F*_1,65_=5.58, *P*=0.021) rather than the unaffected (*F*_1,63_=1.36, *P*=0.247) side with estimated differences of 4 and 2 CA points respectively. Further, in the mid-late time-window, although the main effect of group was not reliable, the difference between HV and CRPS was notably greater in the passive (*F*_1,64_=0.99, *P*=0.324) compared to the active (*F*_1,66_=0.06, *P*=0.802) condition, with patients scoring 1.5 CA points higher on average in the passive, and 0.3 points lower than controls in the active condition. Overall, there was no consistent support for our second hypothesis – H.2 –, stating a larger difference between hands in the CRPS group compared to HV. An expected result, although not formally stated *a priori,* was the effect of attention. The decoders trained on the correct trials from the active condition performed reliably better than the passive ones in the two later time windows (*F*_1,74_ = 29.94, *P* <0.001, *η*^2^_*p*_ = 0.237; F1,74 = 40.552, *P*<0.001, *η*^2^_*p*_ = 0.301). Of note is also a reliable main effect of side on peak latency in early (*F*_1,72_ = 13.60, *P*<0.001, *η*^2^_*p*_ = 0.121) and mid (*F*_1,94_ = 12.39, *P*<0.001, *η*^2^_*p*_ = 0.116) windows with the affected/non-dominant hand peaking on average 5.4 and 8.2 ms earlier than the unaffected/ dominant counterpart. In the mid time-window, CRPS patients had later CA peaks compared to controls (*F*_1,94_ = 7.40, *P* = 0.008, *η*^2^_*p*_ = 0.073).

Importantly however, a different set of results emerged when the classification included also error trials from the active condition. No evidence to suggest a lower mean decoding in the CRPS patients compared to controls was present in any of the four time-windows. Notably instead, a reliable effect of group, in the opposite direction compared to our predictions (i.e., CRPS > HV) emerged in the mid-late time-window of 180-320ms (*F*_1,24_ = 7.37, *P* = 0.012, *η*^2^_*p*_ = 0.095), further corroborated by exploratory analysis of peak performance (*F*_1,24_ = 8.66, *P* = 0.007, *η*^2^_*p*_ = 0.095). Additional analyses performed by excluding early RTs (< 500ms) confirmed this unexpected midlate-window group difference, and further suggested that it may be unlikely explained by motor implementation differences.

### Individual-level results

The subject-wise differences between the two sides (affected and unaffected) in each condition and timeframe are summarised in *Supplementary Figure 3*, and all statistical results (including F-values, p-values and effect sizes) are fully reported in *Supplementary Table 5* (correct trials only) and *Supplementary Table 6* (all active trials).

When only correct identification trials from the active condition are considered, 2 of the 11 patients with sufficient active data showed a better performance in the unaffected compared to the affected side (*F*_1,9_ = 9.45, *P*=0.013, η*²p*=0.512; *F*_1,9_ =6.66, *P*=0.030, η*²p*=0.425), as proposed in H.3. However, one patient also reported a reliable effect of size in the opposite direction with respect to our predictions (i.e., a better performing affected side). None of the 11 patients showed an interaction of side by condition as we hypothesized in H.4. On the contrary, 2 patients had a reliable interaction effect such that the distance between classification performance on the two sides was enhanced by the active condition demands (*F*_1,9_ = 19.01, *P*=0.002, η*²p*=0.679; *F*_1,9_ =7.08, *P*=0.026,η*²p*=0.440).

When error trials were included in the classifier training, we found an effect of side in the predicted direction (H.3) in 3 of 13 patients (*F*_1,9_= 7.80, *P*=0.021, η*²p*=0.464; *F*_1,9_=5.72, *P*=0.040, η*²p*=0.388; *F*_1,9_=7.77, *P*=0.021, η*²p*=0.463). None of the 13 participants affected by CRPS showed interactions as we hypothesized in H.4, however, in 6/13 of the patients’ results showed the “shape” in the predicted direction. An effect of side leaning in the opposite direction with respect to our prediction (i.e., the affected side performing better than the healthy one) was present in two other patients (*F*_1,9_=12.21, *P*=0.007, η*²p*=0.576; *F*_1,9_=5.54, *P*=0.043, η*²p*=0.381), one of which also showed an interaction of side by condition (*F*_1,9_=21.24, *P*=0.001, η*²p*=0.702), but in the opposite direction to our hypothesis H.4.

Overall, there was no reliable effect for our third hypothesis of better classifier’s performance in the unaffected side compared to the painful one, and no support for H.4 (reduction of affected-unaffected gap in the active condition).

### Temporal Generalization and Topographical maps

The Temporal Generalization matrices resulting from our exploratory TGA within the two groups, sides and conditions are represented in *Figure 5*. The active condition outputs (Figure 5B, D and F) are extracted from correct trials only. The first considerable difference that stands out (by contrasting Figures 5A, C and E to Figures 5B, D and Figure 5F) is how information processing changes radically when task demands are introduced in the active condition. While during passive stimulation (5A-C-E) a diagonal-shaped decoding performance indicates neural patterns that generalize over a brief, transient time period only, higher levels of temporal generalization (square pattern) emerge during the active condition (5B-D-F), suggesting cortical signals (or mental representations) that are “maintained” more stable across time. Secondly, when we focus on the active processing differences between sides of the body, i.e. unaffected (5B) and affected (5D) by the CRPS, is noticeable how, while the unaffected side shows a generalization pattern that is comparable to that of healthy volunteers (5F), with a more widespread decodability, the affected side TGA seems to yield a narrower and “delayed” spread of finger neural representation.

**Figure 5.**
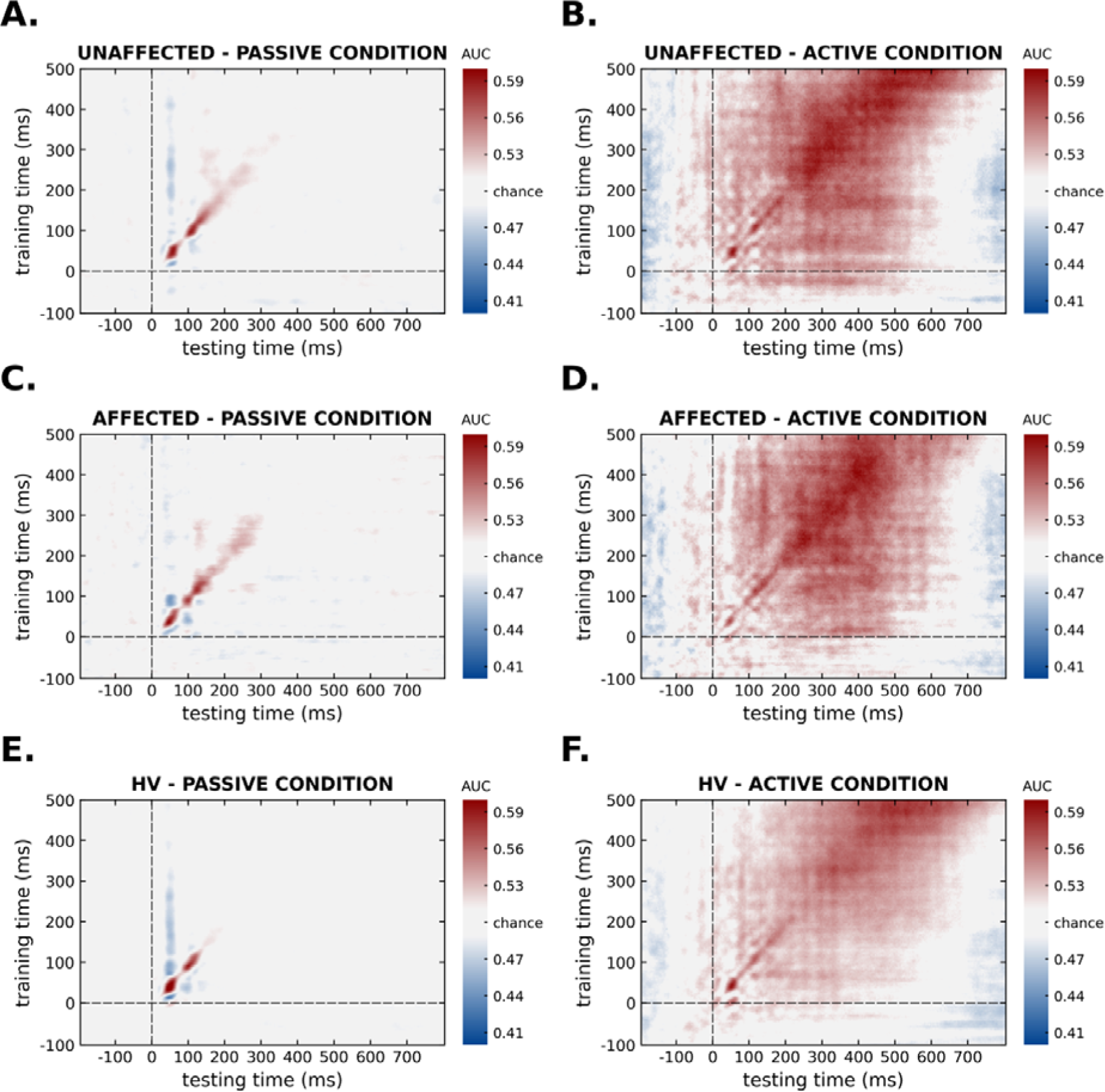
Temporal generalization matrices representing the generalization across time for **(A)** CRPS patients’ unaffected side of body in the passive condition, and **(B)** their unaffected side data in the correct trials from active condition; **(C)** CRPS patients’ side affected by the disease in the passive condition, and **(D)** their affected side data during the active condition (correct trials only). Healthy volunteers’ temporal generalizations are also plotted for comparison in the **(E)** passive condition, and **(F)** in the active condition (correct trials only). In each of the six graphs, training timepoints are represented on the y-axis, while testing timepoints are displayed on the x-axis and two dashed lines locate the time of tactile stimulus onset in both dimensions. Color indicates decoder’s performance strength (i.e., classification accuracy). The diagonal values (from the left bottom to top right) closely correspond to the outcome computed in the previous analysis (represented in Figure 4E, 4F and 4A, 4B). Higher off-diagonal performance indicates stronger temporal generalization (i.e., stability of neural representations).

To better characterize the spatial aspects of these processing differences we represented time-specific topographical maps for all conditions, groups and sides in *Supplementary Figure 5*. Three time-points revealed reliable clustering of electrodes after permutation: 124 ms after stimulus presentation during passive simulation of the unaffected side of CRPS patients (*Supp. Figure* 5B); 460 ms after active stimulation of the left hand of HV (*Supp. Figure* 5C); and 404 ms after active stimulation of the unaffected hand in the CRPS group (*Supp. Figure* 5D).

## Discussion

We confirmed a decreased in finger discrimination performance in CRPS patients compared to healthy participants, following decreased accuracy, slower reaction times and increased interindividual variability (as described here and in Kuttikat et al., 2018). At the neural level we show support for lower early cortical decoding performance in the CRPS group compared to healthy participants (H.1) after tactile stimuli but not enough evidence for a greater decodability gap between hands in these patients in any of our time windows of interest (H.2). In addition, ∼20% of patients showed a reliably worse classification performance on the affected (compared to the unaffected) side of the body (H.3), but none of our patients presented a reliable reduction of decodability gap between hands during the active task, compared to the passive stimulation (H.4).

With the exception of H.1, that was confirmed in the early time-window only when error trials were excluded from classifier training, these conclusions were robust across sensitivity analyses including correct trials only, all active trials or only trials when response was given after 500 ms.

### Study limitations and strengths

Several words of warning feel necessary before delving deeper into the interpretation of this set of results. The first concerns the small sample size, that strongly limits the generalizability and robustness of our analysis. In addition, the experimental protocol adopted for this study was developed with a close eye to minimizing patient discomfort, but it sacrifices certain design considerations. For instance, the passive task was always preceding the active one, and left / right hand stimulations were alternated rather than randomized (*Figure 1*). This fixed condition sequence may have possibly introduced unmeasured, time-dependent fatigue effects that would impact the interpretability of the cognitive effects we highlight. At the same time, the alternating hand stimulation is likely to elicit a lesser spatial attentional demand compared to a randomized alternative. Moreover, because the experiment was interrupted whenever patients reported pain or uncomfortableness, this may have introduced some form of selection bias, especially with respect to levels of allodynia, possibly affecting the clinical representativeness of our sample (*Supp. Table* 1). Several analytical strategies were also adopted in the present work, i.e. *one-vs-all*, *all-vs-all* and *1-vs-5*, each hinging on slightly different assumptions and offering peculiar advantages and disadvantages. Although the purpose of this study was to advocate for the utility of a new technique, and showcase the available analytical options it may offer, future efforts should focus on settling a standard for investigation.

Nonetheless, the adoption of time series MVPA has proven a useful novel tool for the investigation of tactile processing differences induced by CRPS. Because of its excellent temporal resolution, portability and low cost, EEG is a precious tool to investigate possible functional changes that may act as biomarkers of disease. However, classical temporal-spatial ERP analyses and source reconstruction efforts (that constitute the majority of this field of investigation) are not robust against many, often unmeasured inter-individual differences. For example, idiosyncratic anatomical differences in the folding pattern of the cortex, skin conductance, age, medication use and psychopathology are all known confounders and they are all extremely variable within the CRPS population. In this context multivariate analysis can better reflect and leverage individual patterns of activation, and, as it does not require any a-priori electrode selection, it potentially opens a new window to explore more distributed processing across the scalp (Fahrenfort et al., 2018) as well as more spatially variable representations across individuals. In addition, the classifiers used for decoding make use of information in the data that is not available when comparing averaged signals, leading to a much increased sensitivity for detecting differences between experimental groups and conditions. It is important to stress that, because of its novelty, the robustness of this method is yet to be tested. Nevertheless, it is encouraging to notice, for instance, how the “super-subject” decoding time-course did yield similar “peaks” to those often reported in in classical somatosensory ERP research: the earliest cortical components emerging between 20-80ms, followed by a N1-like wave (commonly observed around 80-120 ms) (Luck, 2005) can be appreciated in both conditions (*Figure 3*). These results lend further support to the thesis that CRPS does not show changes in the fine grained cortical somatotopies (Mancini *et al*., 2019). These two early modulations are meant to mirror the more “physiological” aspects of sensory processing, tracking afferent inputs from peripheral nerves into primary cortical areas (S1; <50ms (Lueders *et al*., 1983; Wood *et al*., 1988)), and from there to secondary processing stages (S2; >80ms), building up to the (yet unconscious) detection of sensory stimuli (Schubert *et al*., 2006; Schubert *et al*., 2008; Garrido *et al*., 2009; Chennu *et al*., 2013). Interestingly, when an attentional modulation is introduced by task demands in the active condition (Figure 3B and 3D) two further and wider performance peaks emerge between 250 and 500 ms (Figure 3B), strongly resembling the notorious P3 (a and b) modulation (Polich, 2007).

### A disease-induced “temporal dragging” of sensory information processing?

The amount of neural information detected by our classifiers was reliably reduced in the CRPS group between 28 and 56 ms after the touch has been delivered. This was true for the passive as well as the active condition (Figure 4 A and B), but, although a similar CRPS-related dampening of early performance peaks was present across sensitivity analyses (Supp. Figure 2), only when error trials were excluded from analysis this effect reached significance. Interestingly, when all trials from the active condition are included, a new group effect emerges, i.e., an abnormally enhanced (rather the reduced) performance in the CRPS group between 180 and 320 ms (Supp. Figure 2A). This arguably “compensatory” shift in the temporal distribution of tactile information processing, i.e., a diminished early performance followed by heavier reliance on later processing, is evident not only in the active condition (Figure 4B), but also to a lesser extent during passive stimulation. Indeed, by looking at the decoding output in Figure 4A and comparing *Supplementary Figure 5* A and B, one can notice how two lower performance peaks in CRPS (vs. HV) round 50 and 100 ms are followed by a “dragging” of decoding sustained over 200-250 ms into the trial, when HV decoder is already at chance. This observation also matches previous evidence of greater amplitude of mid-late ERP components in CRPS patients (Kuttikat *et al*., 2018) and potentially signals further processing or delays of sensory sensation leaking into central cognitive control or awareness processes. The passive “dragging effect” emerges again from TGA (Figure 5A and C vs. Figure 5E), stressing again how in CRPS patients compared to HVs, a shade of relevant finger information does persist beyond basic sensory processing stages, despite no instructions are currently deeming that information useful to fulfil any higher-level task. Altogether, these results are in line with the prior analysis of this dataset (Kuttikat *et al*., 2018) as well as other recent theoretical accounts (Brown *et al*., 2020), suggesting that later (“P2 / P3-like”) components of processing, supporting higher cognitive functions such as attentional allocation and perceptual decision-making, assume greater importance in CRPS patients compared to controls and we argue that these later-latency, more “cognitive” stages of processing, ultimately may be the ones that enshrine the key to perceptual differences induced by chronic CRPS.

### An affected-side-specific “temporal shift” into later processing stages

Although the effect of side as we expected it (affected < unaffected) did not reach a robust statistical support neither at the group nor at an individual level, when zooming into the decodability time-courses corresponding to the affected and unaffected sides of the body (Figure 4E and 3F) some interesting patterns emerge. In the passive condition for instance (Figure 4E), one could notice how the above mentioned “dragging” of performance around 200-250 ms in the CRPS group (Figure 4A; Supp. Figure 5B) seems to be mostly driven by the affected side, rather than by the unaffected counterpart. Although this is a strictly qualitative interpretation, and hence speculative, we suggest this affected-side-specific docking of neural information onto a time-frame that is thought to underlie more cognitive stages of processing (rather than early perceptual/sensory), could imply that more advanced / abstract aspects of perception are altered by the disease, especially on the side of the body that is directly affected by it. Similarly, in the active condition, one could speculate how the CRPS-related reduction of the first peak in Figure 4B (50 ms, supposedly underlying poorer primary somatosensory processing, and mirrored in the passive condition in Figure 4A) seems to originate again slightly more from the affected rather than the unaffected side of the body (Figure 4F and Supp. Figure 5D). Interestingly, the exploratory analysis of the temporal dynamics of processing in the affected and unaffected sides of CRPS patients (Figure 5B and 5D) helped highlighting a considerable temporal shift in finger representation that seems specific to the affected side, but could not emerge by simple diagonal (i.e., transient) decodability (Figure 4 and 5). More research is needed to clarify these side-specific dynamics of perceptual information processing in CRPS, but here we highlight a potentially crucial role of attentional demands / task instructions in such distinction (Bultitude *et al*., 2017).

### Intersubject variability: an issue and a resource

Besides the underappreciated role of later stages of processing in CRPS-related somatosensory misperceptions, the second most consistent result emerging from our analysis is that of a substantial inter-subject variability in the CRPS group. We snapshot this variability in *Supplementary Figure 3*, by representing the mean decoding difference between the two hands (i.e., “decodability imbalance”) in each participant, condition, and time-window. There, while healthy volunteers (Supp. Figure 3B) are more consistent in showing a slightly better performance on the dominant hand across conditions and time-frames (with higher imbalances early in the trial quickly resolved after 80ms), CRPS patients show less overlap between conditions and a less consistent pattern across the group and across time. Interestingly, behavioral results point again to a more variable picture in the CRPS group compared to HVs. Even though patients did show lower sensitivity (*d’*) scores than controls on average, most of the variability in *d’* scores was due to interindividual variability rather than any of the fixed effects in the model, and by inspecting Figure 2C, one can appreciate how the spread of *d’* values is relatively larger in the CRPS group compared to controls.

Similarly, while the expected negative criterion shift (Bang & Rahnev, 2017) is consistently present across healthy participants, CRPS patients’ decisional process seems much less consistent (Figure 2B and 2D) and overall less finely tuned to statistical optimality (Brown *et al*., 2020). In light of this issue, future analyses oriented towards leveraging rather than silencing these inter-individual differences, can greatly contribute to our understanding of misperception phenomena in CRPS.

### Theoretical interpretation: Hierarchical Predictive Coding

The journey of CRPS through possible theoretical explanations and framings has been a troubled one (Feliu & Edwards, 2010; Borchers & Gershwin, 2014; Popkirov *et al*., 2019). Most recently, some authors began to appeal to the idea that, in order to understand the complex and subtle perceptual phenomena accompanying the disease, an articulated and thorough model of human perception needs to be taken into account (Kuttikat *et al*., 2016; Kuttikat *et al*., 2018; Popkirov *et al*., 2019; Brown *et al*., 2020). If we model the human brain (including that of CRPS patients) under a framework that regards its mechanisms as a complex, hierarchical – possibly Bayesian – structure, we can then assume perception is simultaneously and dynamically influenced not only by afferent inputs and forward sensory processing steps but also by higher-level knowledge. In a nutshell, this long-standing perspective, often referred to as Hierarchical Predictive Coding (HPC) models (Friston, 2018), proposes that information accumulated and distilled from prior experiences is constantly used to predict or “explain away” new sensory inputs; if the new experience is completely conforming to such expectations, no further processing is needed, but when something surprising (i.e., not matching the existing prediction) is present in the incoming sensory stream, a residual prediction error is generated. These errors propagate forward in the system and are used to update the expectations and improve future predictions, i.e., learning (Feldman & Friston, 2010). Two aspects of HPC theories are relevant when trying to understand tactile misperceptions in CRPS. One hypothesis is that ambiguity in sensory input (resulting for instance from sensory nerve pathology in some forms of CRPS) biases perception towards expectations, implying an abnormally strong influence of cognitive predictions on perception. Some evidence pointing in this direction is that lower frequency bands (e.g., delta and theta ranges) of brain activity, thought to underlie top-down predictions, show greater spectral power across frontal and somatosensory cortices in CRPS patients compared to controls (Walton *et al*., 2010). The same idea also finds support in our group-level decoding output (Figure 4A and 4B), where an initial lesser decodability in the CRPS (vs. HV) in the first (50ms) peaks, is inverted in later time frames (after 180ms; our mid-late time-window). Zooming into the active identification task, a heavier reliance on expectancy-related information could be a plausible explanation of the group gain in classifier performance (Figure 4B) as well as the affected-side specific delay in establishing a stable finger mental representation to support digit identification (Figure 5D). This “shift” of processing (described above) could be interpreted as a “compensatory” mechanism, that leverages the intact top-down neural structure to a newly lesioned and inefficient afferent stream, but, an internal model of the world that is less efficient to the incoming information ultimately becomes too rigid and bound to miss the optimal interpretation of such sensory input. Indeed, our behavioral exploration showed a much less uniform estimated displacement of decision criterion (*c*; Figure 2B and 2D) in the patient group, and this was especially true for the affected side of the body. CRPS patients’ perceptual decisions were less tuned towards the optimal value with respect to the real probability of stimulation (Bang & Rahnev, 2017) and this idea echoes with other similar but independent analyses of CRPS-related changes in perceptual predictions (Brown *et al*., 2020). Altogether, the picture that seems to emerge is that of a lack of balance between top-down and bottom-up influences on perception, which brings us to the other key player in this complex Bayesian game of perception: attention. In the HPC framework, attention (or “salience”) serves as a balance-control system, enhancing or dampening the influence of the other model components (bottom-up or top-down signals) according to the respective “precision” weights, i.e., degree of certainty (Chennu *et al*., 2013; Moran *et al*., 2013). This could be relevant to CRPS research, for two reasons. First, pain (especially chronic pain) is known to drain attentional resources from other parallel processes (Dick *et al*., 2003) thereby impairing identification performance on the affected, but also potentially on the unaffected side.

Second, a lack of attention (or precision) on the affected side could prevail in patients with more severe cognitive neglect-like symptoms (Kuttikat *et al*., 2016; Wittayer *et al*., 2018), resulting in worse top-down/bottom-up balancing on the affected rather than the unaffected side. This potentially concurring antagonist mechanisms could explain why, despite an affected-side specific delay in processing was evident from TGA outcomes (Figure 5B-D) the difference between sides in terms of behavioral (Figure 2) and diagonal decoding performances (Figure 5F) was less clear and inconsistent across the patient group, as emerged from the individual-level statistical results as well. Finally, one interesting result emerging across all analyses is the lack of detected differences between upper and lower limb CRPS patients. Although we did not have the power to formally test these differences, in our limited sample, both the neural and the behavioral performance were similar regardless of the area (upper or lower limb) affected (Supp. Figure 4). We interpreted this unexpected result as a further hint that abnormal processing in CRPS may be driven by higher-order alterations rather than primary sensory processing impairments.

### Alternative interpretations: motor planning and implementation?

Of course, attentional allocation and decision-making are not the sole “mid / late” processes capable of driving neural patterns, and thereby decoding performance, during the active task. One important objective of future research would be for instance to distinguish perceptual processes (i.e., finger detection, recognition and attention) from the motor planning and its implementation (i.e., finger ID’s pronunciation). In our sample, the motor response for all CRPS patients and the majority of the healthy volunteers took place around 1000-1500 ms after stimulus presentation, whereas the estimated motor planning timing ranges around 300-400 ms before movement execution (Shibasaki & Hallett, 2006). Moreover, the sensitivity analysis we conducted by excluding all early response (< 500 ms after stimulus presentation) trials, largely mirrored the patters detected in the full set analysis and suggest that the late processing differences detected by the classifier reflect perceptual rather than premotor or response-related alterations. Some weak evidence against a premotor explanation of detected differences comes from the observation of scalp distribution of decoder weights (*Supplementary Figure 5*) and RTs (*Supplementary Table 2*). Particularly, in graphs C and D of *Supp. Figure 5*, it’s noticeable how the spatial patterns generating later peaks of performance are not homogeneous across time, and they do not overlap with the expected distribution underlying motor preparation or planning (Jagannathan *et al*., 2021). However, it is important to keep in mind that the small sample size and the suboptimal nature of our task (from a cortical differentiation perspective) may have heavily hindered the reliability of these spatial clustering results.

### Conclusion

Altogether, the puzzle of symptoms and neural information patterns characterizing this enigmatic condition seems to point to a difficulty with the body mental representation (Kuttikat *et al*., 2016; Halicka *et al*., 2020), rather than a mere deficit in peripheral transmission (Yvon *et al*., 2018) or modifications of cortical somatotopies (Mancini *et al*., 2019). Expectancy-related and attentional aspects of sensory processing deserve more investigation as contributing mechanisms to CRPS perceptual disturbances, a combination of novel decoding techniques and computational models such as HPC may be useful for understanding the cognitive and pathophysiological changes related to them. More research is needed to clarify side-specific and general dynamics of perceptual information processing in CRPS, ideally supported by principled experimental design and guided by the ultimate goal to leverage individual differences to boost understanding, prevention and treatment of the disorder.

### Author Contributions

TB and NS were responsible for the conceptualisation and preparation of study protocol. Anoop Kuttikat recruited the subjects, administered the tests and collected all EEG data. CB provided the data and, together with TB, NS, and MN offered vital feedback on analysis planning and interpretation of the results. SD and TB prepared the preregistration and SD analysed the data. SD, MN, NS, CB and TB wrote the paper based on SD Master Thesis.

### Data Accessibility Statement

All data supporting the findings of this study are available upon request to CB. The data are not publicly available as they contain sensitive information that could compromise the privacy of research participants (patients). All script and analyses can be accessed directly (https://github.com/SereDef/CRPS-decoding-behavioral-modeling).

## Acknowledgements

This research was funded by BJA/RCoA (Project Grant ID WKR0-2018-0060 to TAB, MN, NS and CB) and Cambridge Arthritis Research Endeavour (CARE to NS). The current analysis was conducted thanks to Erasmus + for traineeship funds to SD. This paper is part of SD master thesis.

## Abbreviations

BDM: Backward Decoding Model
CA: Classification Accuracy
CRPS: Complex Regional Pain Syndrome
EEG: Electroencephalography
ERP: Evoked Related Potential
HPC: Hierarchical Predictive Coding
HV: Healthy Volunteers
MVPA: Multivariate Pattern Analysis
RCR: Random Classification Rate
RT: Reaction Time
S1: Primary Somatosensory Cortex
S2: Secondary Somatosensory Cortex
TGA: Temporal Generalization Analysis.

